# Understanding Timing of Autism Diagnosis: Impact of Sociodemographic Factors, Verbal Ability, and Sex

**DOI:** 10.64898/2026.06.01.26354604

**Authors:** Allison Jack, Jessica V. Smith, Goldie McQuaid, Lauren Kenworthy, Alexis Khuu, John F. Strang, Gregory L. Wallace, Allison B. Ratto

## Abstract

**Background:** Female individuals tend to be diagnosed with autism later. One factor suggested to contribute to diagnostic timing is verbal ability, in which autistic females may show strengths relative to male peers. Social drivers of health (SDOH) predict higher verbal skills, yet access to resources may facilitate diagnosis; thus, SDOH likely contributes to diagnostic timing in complex ways. We use data from two autism cohorts with substantial representation of those assigned female at birth (AFAB) to examine interactions among assigned sex at birth (sex), verbal IQ (VIQ), and SDOH in predicting autism diagnostic timing.

**Methods:** We used multiple linear regression to examine sex assigned at birth and VIQ as predictors of diagnostic timing in an assigned-sex-balanced *research sample* (*N*=164, AFAB: 71) and an independent *clinical sample* (*N*=641, AFAB: 177). We hypothesized VIQ would positively predict diagnostic age, particularly among AFAB. Available data in the clinical sample also permitted us to explore the contributions of SDOH and inclusion criteria to model fit in this cohort.

**Results:** In the *research sample,* VIQ, but not sex, positively predicted diagnostic age. In the *clinical sample*, VIQ and VIQ×SDOH, but not sex, predicted diagnostic age. Fitting the same model in a *subsample* of the clinical cohort formed by applying exclusion criteria used in the research sample (*N*=484, AFAB: 110), VIQ×SDOH×Sex became significant. For AFAB, higher VIQ and lower SDOH together were associated with later diagnosis in the clinical subsample, while for AMAB the opposite was true.

**Conclusions:** Autistic youth with strong verbal ability may experience diagnostic delays. SDOH interacts with VIQ in a complex fashion, with lower SDOH generally exacerbating the tendency for VIQ to be associated with later diagnosis across a large clinical sample. However, among autistic youth without complicating medical factors or intellectual disability, this relationship is dependent upon sex.

---

Autism is identified at a significantly higher rate in male than female individuals, with most large epidemiological surveys reporting a ∼3.4:1 ratio (Shaw et al., 2025). However, a growing body of research suggests that observed prevalence ratios are partly driven by systemic sex/gender bias, leading to under-recognition of autism in female individuals. Of note, most autism research to date has not distinguished between assigned sex at birth and gender, although gender identity is distinct from assigned sex at birth, and gender diversity is more prevalent in autistic^1^ than in neurotypical samples (Dewinter et al., 2017; Walsh et al., 2018; Warrier et al., 2020). In this background section, we use the term “sex/gender” to denote the lack of distinction between these constructs in most extant literature; further, while we describe literature on autistic “girls/women,” we note people referred to as “girls” and “women” in the literature cannot be assumed to identify as such in work where gender identity is not assessed.

Studies investigating potential sex/gender bias in autism prevalence rates have reported smaller or even equivalent sex/gender ratios in diagnoses and/or social communication profiles, after accounting for methodological differences in ascertainment and sex/gender biases in measurement tools (Burrows et al., 2022; Loomes et al., 2017). There are also indications that girls/women are excluded from autism research at higher rates than boys/men (D’Mello et al., 2022), resulting in samples that are under-powered to investigate sex and gender differences, thus potentially yielding flawed conclusions (Mo et al., 2021).

Girls/women also tend to be diagnosed autistic later than boys/men (Dillon et al., 2021; Fusar-Poli et al., 2022; Kavanaugh et al., 2021; McCormick et al., 2020; McDonnell et al., 2021), with some variability across studies (Davidovitch et al., 2022; Mazurek et al., 2014). Delays in autism diagnosis among girls/women are consistently greatest among those with strong cognitive and verbal abilities (Giarelli et al., 2010; Harrop et al., 2021; Salomone et al., 2016; Shattuck et al., 2009). Notably, diagnostic delays persist even without significant differences in the timing of parental first concerns (Dillon et al., 2021; Hiller et al., 2016; McDonnell et al., 2021). Indeed, both retrospective studies of autistic children and prospective work with infant siblings have found generally comparable clinical profiles across sex/gender in the preschool period (Burrows et al., 2022; Davidovitch et al., 2022; Hiller et al., 2016; Little et al., 2017). Thus, these delays seem to be driven by sex/gender disparities in the autism diagnostic process, rather than lack of early signs. Consequently, many autistic girls/women are not diagnosed until adulthood, after several evaluations and other psychiatric diagnoses (Baldwin & Costley, 2016; Bargiela et al., 2016; Fusar-Poli et al., 2022; Geurts & Jansen, 2012).

Under-recognition of autism and delays in identification among girls/women has been theorized to be partially attributable to differences in the behavioral manifestation of autism by sex and gender. Clinicians may be less likely to consider an autism diagnosis for girls/women (Baldwin & Costley, 2016; Whitlock et al., 2020) and perceive autism identification to be more challenging in girls/women (Jamison et al., 2017; Tsirgiotis et al., 2022). Among individuals meeting gold-standard, research criteria for autism, both meta-analytic studies (Saure et al., 2023; van Wijngaarden-Cremers et al., 2014) and large-scale multi-site investigations (Kaat et al., 2020) generally find little to no evidence of sex/gender differences in social-communication skills, though slightly lower rates of restricted/repetitive behavior and interest (RRBI) symptoms in girls/women. However, there is growing evidence of subtle sex/gender differences in phenotypic presentation of autistic traits, primarily in autistic individuals without co-occurring intellectual disability (Lai et al., 2022; Saure et al., 2023). This is notable, as higher intellectual abilities are also associated with delays in age of diagnosis (Daniels & Mandell, 2014; Mazurek et al., 2014; McDonnell et al., 2021) with more advanced verbal abilities being particularly associated with later diagnostic age (Harrop et al., 2021; Salomone et al., 2016), and some of these studies finding that effects are particularly pronounced among girls.

Autistic girls/women may have better language skills on average than their male peers, particularly in pragmatic communication and narrative language (Conlon et al., 2019; Parish-Morris et al., 2017; Sturrock et al., 2020). They may also show more neurotypical patterns with respect to gestures, facial expressions, and play (Kirkovski et al., 2013; Knickmeyer et al., 2008; Rea et al., 2022; Rynkiewicz et al., 2016; Tsirgiotis et al., 2022), such that they are perceived as having better reciprocal interaction skills (Head et al., 2014; Kanfiszer et al., 2017; Kauschke et al., 2016; Rynkiewicz et al., 2016; Sedgewick et al., 2016; Tsirgiotis et al., 2022). These differences in social presentation have been discussed in the context of camouflaging – the capacity of some autistic people to “mask” their autistic traits by behaving more like neurotypical people (Hull et al., 2017), which may contribute to delays in diagnosis (Belcher et al., 2022). Although camouflaging occurs across sexes and genders, autistic girls/women and female individuals appear to engage in it at a higher rate than their male counterparts (Cook et al., 2021; Lai et al., 2017; McQuaid et al., 2022).

Autistic girls/women may also be less likely to meet diagnostic criteria on gold-standard autism diagnostic measures, perhaps partially due to camouflaging, even when they meet clinical criteria (DSM or ICD) for autism (Dworzynski et al., 2012; Ratto et al., 2018; Wilson et al., 2016). This pattern has been directly linked to the exclusion of girls/women from autism research studies, meaning that as new knowledge of autism is published, it is not based on representative samples of autistic people across sexes and genders (D’Mello et al., 2022). Moreover, delays in diagnosis and consequently, access to support, have been associated with poorer long-term outcomes (Clark et al., 2018; Gabbay-Dizdar et al., 2022) and increased risk for psychiatric difficulties (Hosozawa et al., 2021) across sexes/genders. Delayed identification also eclipses the opportunity to form a positive autistic self-identity early in life, which can exert protective effects on self-esteem and wellbeing (Oredipe et al., 2023). These risks are exacerbated for individuals from households with lower socioeconomic status and/or communities with fewer resources for support of positive child development.

Lower socioeconomic status (a construct typically encompassing individual or household-level factors related to income, educational attainment, and class and/or occupation) has historically been associated with autism diagnostic delays and under-diagnosis (Durkin et al., 2017; Smith et al., 2020), though there are some indications that these gaps may be decreasing (Maenner et al., 2023). At the same time, higher socioeconomic status is associated with higher verbal and intellectual abilities (Noble et al., 2007), and as previously noted, these are associated later age of diagnosis. Increasingly, social drivers (or “determinants”) of health (SDOH) are also being considered as important contextual factors in shaping mental, physical, and behavioral health outcomes. While strongly related to socioeconomic status (Braveman & Gottlieb, 2014), SDOH provide a more multidimensional view on an individual’s health-relevant context by integrating information about community-level resources such as, e.g., access to high-quality healthcare, schools, and clean water (World Health Organization, 2025). Previously, children living in higher-resource neighborhoods have been found to score higher on verbal IQ (VIQ) measures (Loblein et al., 2026), and it has been suggested that SDOH could help explain gender-related patterns of autism underdiagnosis (Hotez & Shea, 2023). Consequently, SDOH and intellectual and verbal abilities are likely to interact and contribute to diagnostic timing in complex ways. Thus, it is critical to evaluate the effects of SDOH, VIQ, and assigned sex on diagnostic timing, as well as their interactions with one another, to understand how this process unfolds in the real world.

This study aims to extend prior work on diagnostic timing by leveraging two samples of autistic youth without co-occurring intellectual disability with differences in case ascertainment approach and sociodemographic composition: 1) a research sample of previously diagnosed autistic youth participating in a study using a cohort balanced by assigned sex at birth and 2) a clinical sample of youth presenting for a first-time diagnosis of autism at a tertiary care clinic in a major metropolitan area. We hypothesized that higher VIQ would be associated with later age at autism diagnosis across samples, and that this risk would be exacerbated among those assigned female at birth (AFAB). We also explored the interactive effects of SDOH with these factors on age at diagnosis. We expected that relationships among assigned sex, VIQ, and diagnostic age would vary depending on SDOH, but given little prior research in this area, made no directional predictions.

## Methods

### Participants

#### Research sample

The research sample included autistic participants for whom autism diagnostic age was available within the first wave of the assigned sex-balanced GENDAAR cohort (NDA Data Collection #2021, *N*=164, AFAB *n*=71). These youth (*M[SD]=*12.50[2.88] years [y]; Range=8.00-17.92y) received diagnoses (diagnostic age *M[SD]*=5.84[3.58]y; Range=1.33-17.00y) prior to recruitment, often through the clinical arm of recruiting institutions. Diagnoses were confirmed with DSM-5 (American Psychiatric Association, 2013) criteria at the time of research participation by expert clinicians using the Autism Diagnostic Observation Schedule 2^nd^ Edition (ADOS-2; Lord et al., 2012) and the Autism Diagnostic Interview – Revised (ADI-R; Rutter et al., 2003). 100% of participants met criteria on ADI-R; one participant did not meet ADOS-2 cutoff score but had a well-established diagnosis through the affiliated clinic. Sex assigned at birth was collected via caregiver report; no assessment of gender was obtained from the cohort at this timepoint, a limitation of this study. Thus, participants are designated *assigned female at birth (AFAB)* or *assigned male at birth (AMAB)*. See Table 1 and Figure 1 for descriptive and distributional characteristics of the research sample.

**Figure 1.**
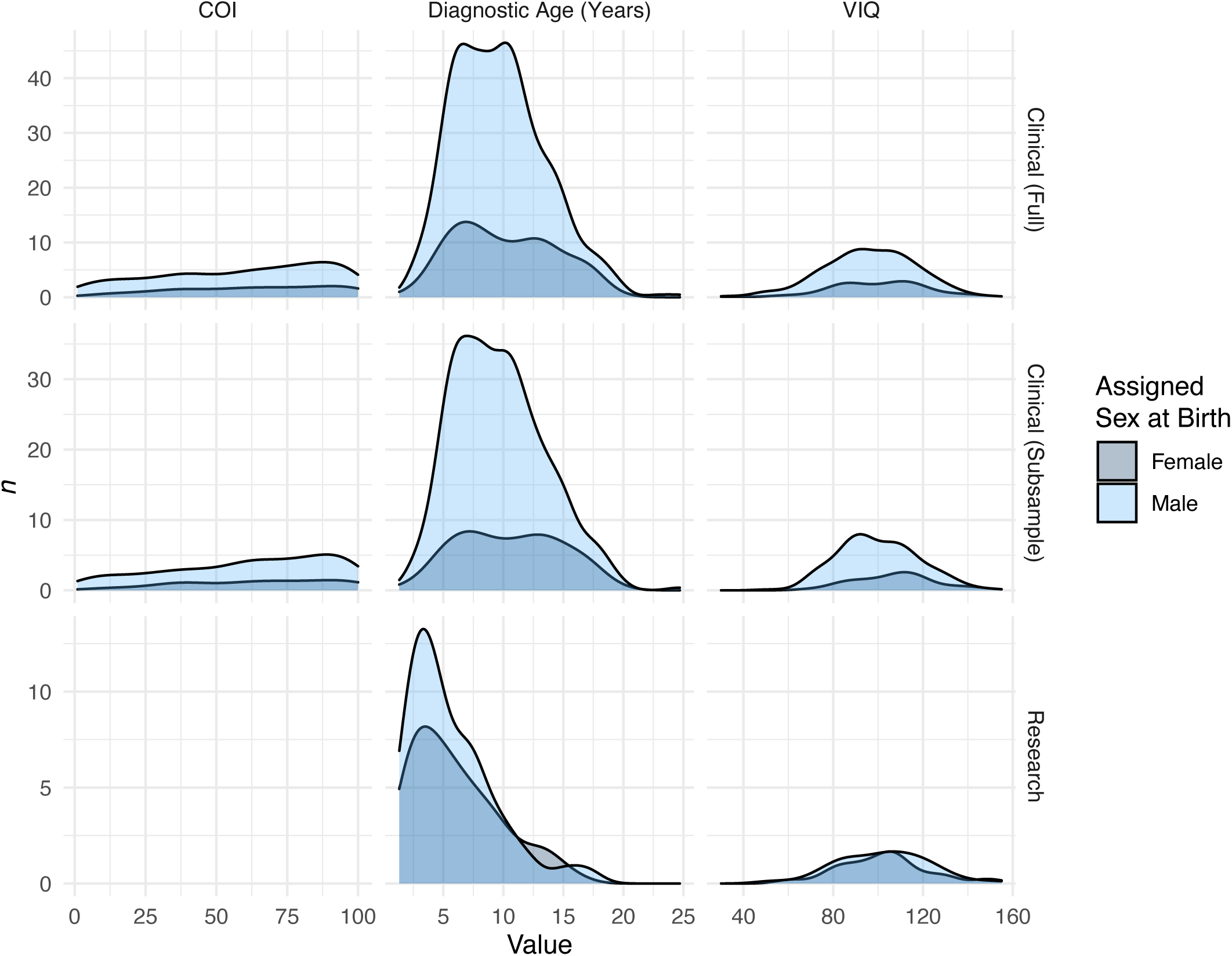
Distributions of continuous predictor and outcome variables by sex assigned at birth, for each sample. Variable distributions for overall Childhood Opportunity Index (COI) scores, Verbal IQ (VIQ) scores, and age of autism diagnosis in years are plotted are plotted as curves with frequency count on the y axis and variable value on the x axis. Fill under the curve indicates assigned sex at birth (female: dark grey; male: light blue). Each variable is assigned a column and each sample (full clinical sample, clinical subsample, and research sample) is assigned a row in the plot matrix.

**Table 1.** Descriptive characteristics of the research sample, by assigned sex at birth.

| Research Sample (N=164) |  |  |  |  |  |  |
| --- | --- | --- | --- | --- | --- | --- |
|  | Female<br>*n*=71 |  | Male<br>*n*=93 |  | Sex<br>Diff. |  |
| | *n* | | *n* | | $\chi^2$ | *p* |
| **Ethno-racial identity** |  |  |  |  |  |  |
| Asian American/Pacific Islander | 2 |  | 1 |  | -- | -- |
| Black/African American | 2 |  | 2 |  | -- | -- |
| Hispanic/Latino,a,e | 7 |  | 11 |  | 0.16 | .803 |
| Mixed | 8 |  | **24** |  | 5.42 | .026* |
| Other/Unknown | 2 |  | 0 |  | -- | -- |
| White | 50 |  | 55 |  | 2.23 | .138 |
|  | *M*(*SD*) | Range | *M*(*SD*) | Range | *t*(*df*) | *p* |
| Age at testing (years) | 12.48(2.87) | 8.0-17.9 | 12.51(2.91) | 8.0-17.9 | -0.07(162) | .943 |
| Age at diagnosis (years) | 6.10(3.67) | 1.3-16.0 | 5.64(3.52) | 1.5-17.0 | 0.81(162) | .418 |
| DAS-II Verbal | 102.42(20.14) | 59-155 | 102.24(20.36) | 52-155 | 0.06(162) | .954 |
| DAS-II Nonverbal | 100.82(17.88) | 67-158 | 102.88(17.08) | 71-149 | -0.75(162) | .453 |
| DAS-II General Conceptual Ability | 100.86(19.93) | 71-162 | 102.57(18.70) | 71-167 | -0.56(162) | .573 |
| SRS-2 total *t*-score (valid<br>*n*=152) | 76.80(10.00) | 57-98 | 73.01(11.06) | 39-93 | 2.18(150) | .031* |
| ADOS-2 CSS | 6.61(1.86) | 4-10 | 7.34(1.84) | 3-10 | -2.54(162) | .012* |
| ADI-R: Social | 18.69(5.44) | 9-30 | 19.29(5.53) | 8-30 | -0.69(162) | .489 |
| ADI-R: Language | 15.73(4.01) | 6-23 | 16.37(4.43) | 8-26 | -0.94(162) | .347 |
| ADI-R: RRBIs | 5.68(2.66) | 1-12 | 6.51(2.74) | 1-12 | -1.94(162) | .054† |

**Note:** Sex differences in observed counts are calculated using chi-squared tests with *p* value computed by Monte Carlo simulation using 10,000 replicates. Differences in means are calculated using *t*-tests. DAS-II: Differential Ability Scales, 2<sup>nd</sup> Edition. SRS-2: Social Responsiveness Scale 2<sup>nd</sup> Edition. ADOS-2 CSS: Autism Diagnostic Observation Schedule 2<sup>nd</sup> Edition Calibrated Severity Score. ADI-R: Autism Diagnostic Interview-Revised. "Social," "Language," and "RRBIs" scores correspond to the diagnostic algorithm scores for "A: Qualitative Abnormalities in Reciprocal Social Interaction," "B: Verbal: Qualitative Abnormalities in Communication," and "C: Restricted, Repetitive, and Stereotyped Patterns of Behavior." Bold font in an *n* column indicates that the observed count for that sex is significantly higher than the expected count. \*: *p* < .05; † *p* < .10

Due to the research aims of the project, which involved multimodal neuroimaging assessment, an inclusion criterion for the cohort was estimated IQ > 70 on one or more of the Nonverbal, Verbal, or General Conceptual Ability Standard Scores of the Differential Abilities Scales Second Edition (DAS-2; Elliot, 2007). Cohort exclusion criteria included: active tic disorder; active seizures during the past year; current use of any benzodiazepine, barbiturate, or anti-epileptic; or twin status (see Jack et al., 2021 for details). As the focus of the project was idiopathic autism, syndromic cases and those with known or likely environmental or medical causes (e.g., significant pre- or perinatal brain injury, severe nutritional or psychological deprivation) were also excluded. Sensorimotor, visual, or auditory impairment after correction that would preclude valid use of diagnostic instruments or experimental paradigms was also exclusionary. SDOH data for the research sample are limited. Household income and maternal education were elicited from participants, but many families elected not to provide this data (household income missing *n*=61 [37%]; biological mother education missing *n*=13 [8%]; Table S1). Consequently, SDOH factors are not included in models analyzing the research sample.

Procedures were approved and data collected under University of California Los Angeles Institutional Review Board (IRB) #10-000387, Boston Children’s Hospital IRB-P00004852, and Yale University Human Investigation Committee #1206010363. Parents provided informed written consent, and children provided written assent.

#### Clinical sample

The clinical sample was drawn from archival data collected through a continuing study approved and monitored by the Children’s National Institutional Review Board (IRB) #Pro00001320. The archival data was collected from a clinical cohort of >5,000 youth seen between 2014-2020 for clinical evaluation in a combined outpatient neuropsychology clinic and multidisciplinary autism center affiliated with an academic medical center in the Washington, DC metropolitan area. From the total cohort, a *clinical sample* (*N*=641) was identified who received a first-time diagnosis of autism at the time of evaluation, had complete data available on key demographic variables and a score reported on the Verbal index of a standardized measure of cognitive ability (i.e., a VIQ score). As such, age at time of evaluation was also age at diagnosis within the clinical sample (*M*(*SD*)=9.80(3.80), Range=1.8-24.7y), and all measures were collected at time of diagnosis, differing from the research sample. All participants were diagnosed with autism by a clinical psychologist or psychiatrist with expertise in autism, based on DSM-5 diagnostic criteria, utilizing developmental history and direct observation and assessment. Among participants who received the ADOS-2 (Lord et al. 2012) as part of their evaluation (*n*=528; 133 AFAB), 96.9% (94.7% of AFAB participants and 97.7% of those assigned male at birth [AMAB]) met cutoff score criteria. Diagnostic information was entered by the evaluating clinician, including whether this was a new autism diagnosis.

Demographic variables (i.e., child race/ethnicity, gender, sex assigned at birth) were gathered via caregiver and child report. To enable analysis of our hypotheses in a broader, more generalizable sample than the research sample, we minimized exclusion criteria. Participants were not excluded based on cognitive ability (i.e., full scale IQ) or co-occurring medical conditions. For the clinical sample, SDOH was characterized using the Child Opportunity Index 2.0 (COI; Noelke et al., 2020), which uses neighborhood-level indicators of SDOH derived from home address. Childhood Opportunity Indices were not collected for the research sample and are not calculable *a posteriori* due to restrictions on the sharing of potentially identifying information among study sites. See Table 2 and Figure 1 for descriptive and distributional characteristics of the full clinical sample. Table S2 provides additional information about co-occurring medical and neurodevelopmental diagnoses in this sample, COI domain scores, and information about alterations to the standard diagnostic assessment (e.g., use of telemedicine).

**Table 2.**
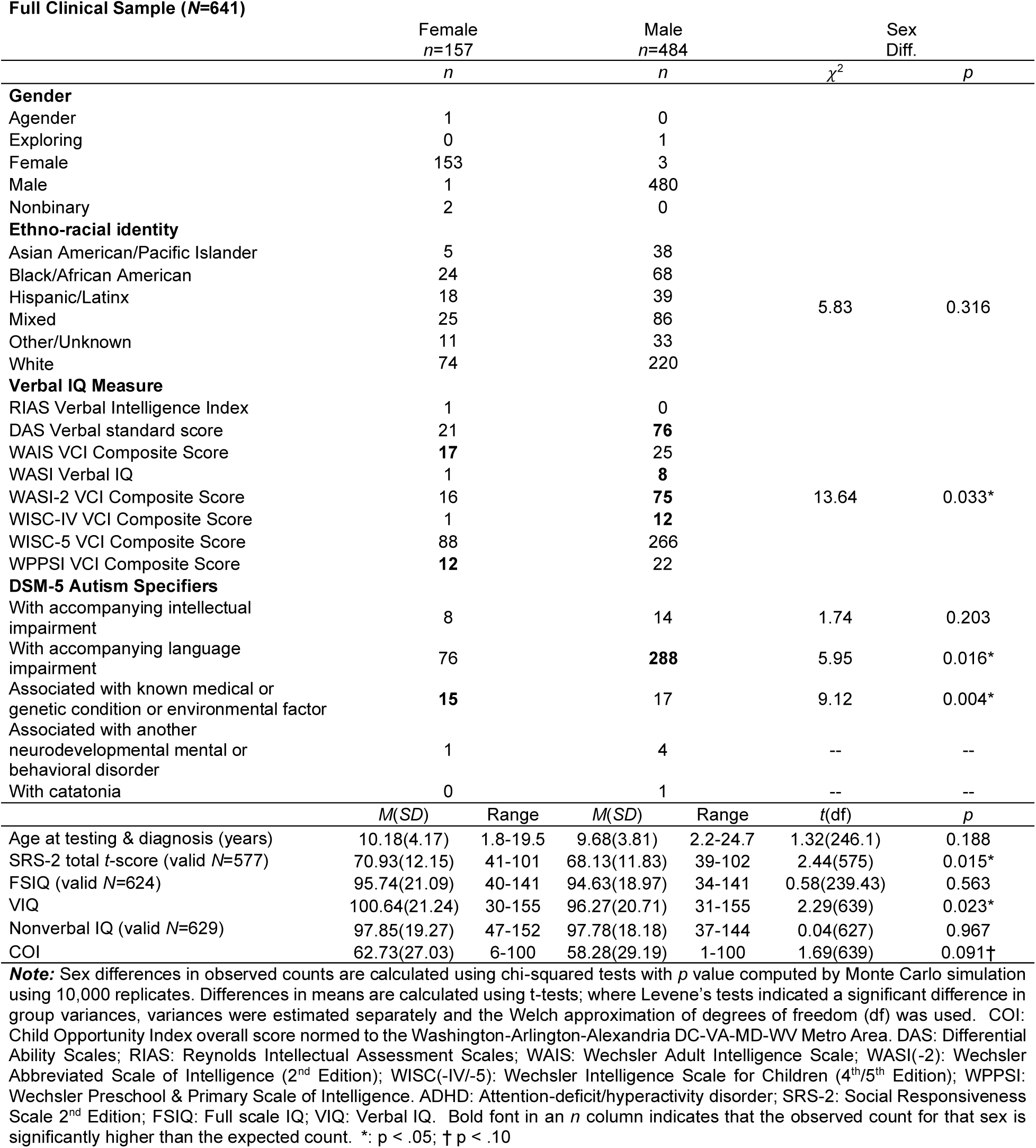
Descriptive characteristics of the full clinical sample, by assigned sex at birth.

To understand how differences in sample inclusion/exclusion criteria potentially impacted our results, we also created a *clinical subsample* that more closely matched the characteristics of the research cohort. This subsample was created by excluding cases from analysis if their reports listed any of the following indicators: non-idiopathic autism; intellectual impairment; intellectual disability diagnosis; FSIQ ≤ 70; or presence of a medical diagnosis or history that would have been exclusionary in the GENDAAR cohort (i.e., brain tumor, prematurity, epilepsy, neurofibromatosis, tic disorder, hydrocephalus, or traumatic brain injury). The resultant subsample consisted of *N*=484 individuals (*n*=110 AFAB; see Table 3 for descriptive characteristics and Table S3 for additional information regarding co-occurring diagnoses, COI domain scores, and assessment adaptations). Analyses were then re-run as for the full sample.

**Table 3.**
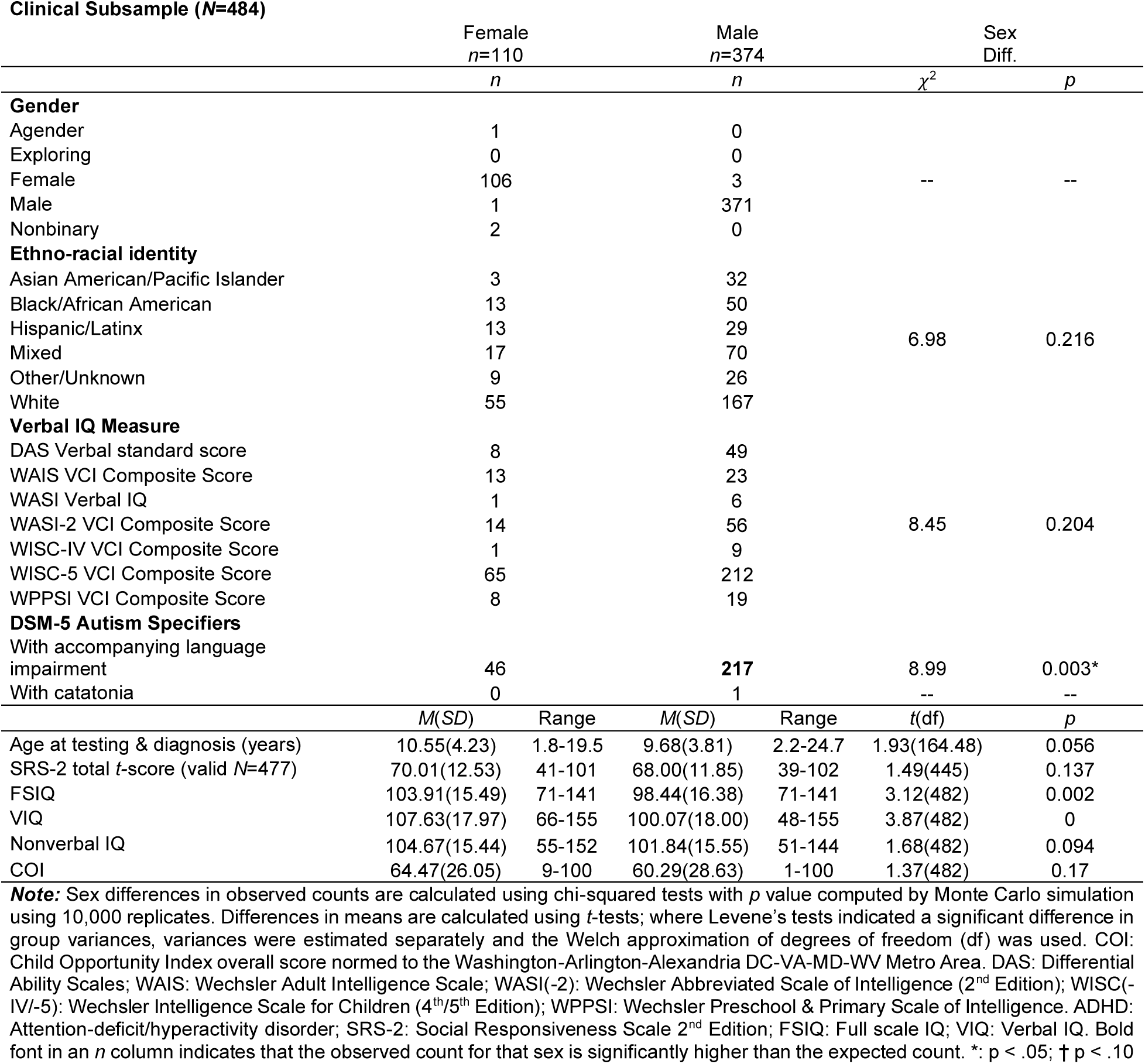
Descriptive characteristics of the clinical subsample, by assigned sex at birth.

### Measures

#### VIQ

VIQ was estimated in all members of the research sample with the DAS-2 Verbal Standard Score (Table 1). In the clinical sample, the instrument used to assess VIQ varied according to clinical judgment, based on the needs and characteristics of the patient (Tables 2-3). The most frequently used measure in the full clinical sample was the Wechsler Intelligence Scale for Children (WISC-5; Wechsler, 2014) Verbal Comprehension Index Score (VCI; *n*=354), followed by the DAS-II (Elliot, 2007) Verbal Standard Score (*n*=97) and the Wechsler Abbreviated Scales of Intelligence 2^nd^ Edition (WASI-II; Wechsler, 2011) VCI (*n*=91). Of note, not all patients were administered assessments in such a way as to produce full scale IQ and/or nonverbal IQ scores, depending on clinical profile and needs (Table 2). Prior research has found no significant differences between VIQ as measured by the DAS-II and the Wechsler scales in autistic youth (Kuriakose, 2014).

#### Child Opportunity Inde

The Child Opportunity Index 2.0 (COI; Noelke et al., 2020) measures the quality of resources and conditions associated with childhood health outcomes using neighborhood-level data. The COI indexes 29 indicators of neighborhood-level data, assessing factors related to Education, Health and Environment, and Social and Economic components. COI scores are generated within each component area, as well as an Overall COI. These scores are then normed (*z*-scores) using national and metropolitan-area norms, which includes the metropolitan center as well as surrounding suburban areas. For analyses in major metropolitan areas, the use of the metropolitan-area norms, rather than national norms, is recommended, as the national norms tend to obscure inequities within metropolitan areas. Thus, COI overall scores normed for the Washington-Arlington-Alexandria DC-VA-MD-WV Metro Area were used, and patients with a home address outside this metropolitan area were excluded from analysis. See Tables S2-S3 for sample descriptives of domain scores (Health & Environment, Education, Social & Economic).

### Analyses

Analyses were conducted in R v.4.2.0; code to replicate these analyses is shared at: [OSF project private until publication]. Hypotheses were tested using linear models fitted using lm in R’s base stats package. Prior to estimation of effects and statistical inference, global validation of linear model assumptions was conducted using gvlma v.1.0.0.3 (Pena & Slate, 2006). When violations of model assumptions were detected, bestNormalize v.1.8.3 (Peterson, 2021) was used to find the variable transformation that best approximated a normal distribution for each variable (e.g., standardized Yeo-Johnson, standardized square root) and the model was re-fit and assumptions re-tested using the transformed data (see Supplementary Information). All final models used normalized variables, passed global validation of linear model assumptions, and had acceptable generalized variance inflation factors (Fox & Weisberg, 2019).

AMAB served as the reference level (AMAB=0) for assigned sex for all models. Significant interactions were probed using the Johnson-Neyman (Johnson & Neyman, 1936) technique. One difference between the research and clinical sample is the timing of diagnosis and assessment of VIQ, as these were concurrent in the clinical sample but not the research sample. To address this difference, a covariate of age at research participation (i.e., interview age) was included in models using the research sample.

Model specification in the research sample was:

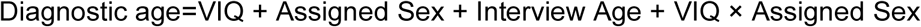

Model specification in the clinical cohort, both the full sample (*N*=641) and subsample (*N*=484) was:

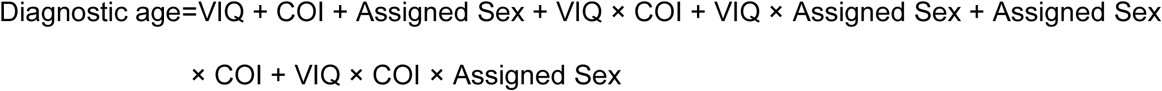

Significance tests for main effect of VIQ were one-tailed, reflecting our *a priori* hypothesis that higher VIQ would predict later diagnosis (that is, have a positive coefficient). Tests for other main effects and interaction terms were two-tailed (non-directional). The Johnson-Neyman technique, or floodlight analysis, mathematically computes regions of significance, or specific value ranges of the moderator at which points the conditional effect of the moderator is significant (Hayes & Little, 2022; Johnson & Fay, 1950).

## Results

### Research Sample

The overall model was significant (*F*(4, 159)=6.28, *p* < .001, Adj. *R*^2^=0.11). The interaction term for VIQ × Assigned Sex was nonsignificant (see Table 4). The only significant main effect was interview age (*B=*0.08, *p* =.002). After removing the nonsignificant interaction term from the model, the effect of VIQ on diagnostic age was significant (*B=*0.24, *p=*0.001).

**Table 4.** Model estimates in research sample.

| Coefficients | $\beta$ | B | SE | *t* | *p* * |
| --- | --- | --- | --- | --- | --- |
| A. Full a priori model |  |  |  |  |  |
| *Reference sex: male* |  |  |  |  |  |
| Intercept | -- | -1.07 | 0.34 | -3.16 | 0.002* |
| Interview Age | 0.24 | 0.08 | 0.03 | 3.15 | 0.002* |
| VIQ | 0.17 | 0.17 | 0.10 | 1.77 | 0.078† |
| Female | 0.06 | 0.11 | 0.15 | 0.76 | 0.446 |
| VIQ × Female | 0.11 | 0.17 | 0.15 | 1.10 | 0.272 |
| RMSE=0.93; Adj. $R^2$=0.16; $F(3,159)$=6.28, $p < .001$ | | | | | |
| B. Model with non-significant interaction term removed |  |  |  |  |  |
| *Reference sex: male* |  |  |  |  |  |
| Intercept | -- | -1.11 | 0.34 | -3.32 | 0.001* |
| Interview Age | 0.25 | 0.09 | 0.03 | 3.32 | 0.001* |
| VIQ | 0.24 | 0.24 | 0.07 | 3.24 | 0.001* |
| Female | 0.06 | 0.11 | 0.15 | 0.77 | 0.444 |
| RMSE=0.93; Adj $R^2$=0.11; $F(3, 160)$=7.96, $p < .001$ | | | | | |

**Note.** \*Significance tests for main effect of VIQ were one-tailed, reflecting our *a priori* directional hypothesis that VIQ would have a positive coefficient. Tests for other main effects and interaction terms were two-tailed (non-directional). $\beta$ : standardized beta. B: Unstandardized beta. \*: <.05 †: <.10

### Full Clinical Sample

VIQ was significantly correlated with COI overall score (*r*(639)=0.24, *p* < .001); however, generalized variance inflation factors for the model were all 1. The overall model was significant (*F*(7,633)=3.57, *p* < .001; Adjusted *R*^2^=0.03; Table 5). Assigned sex and two-way interaction terms including assigned sex were not significant; the three-way interaction among assigned sex, COI, and VIQ was trending (*B*=-0.17, *p*=.062). The simple effect of VIQ *(B=*0.14, *p*=.003) and the two-way interaction between VIQ and COI were significant (*B*=0.10, *p*=.031). A Johnson-Neyman test indicated that the linear estimate of the relationship between VIQ and diagnostic age was significantly greater than zero when (normed; standardized) COI ≥ -0.42 (Figure 2).

**Figure 2.**
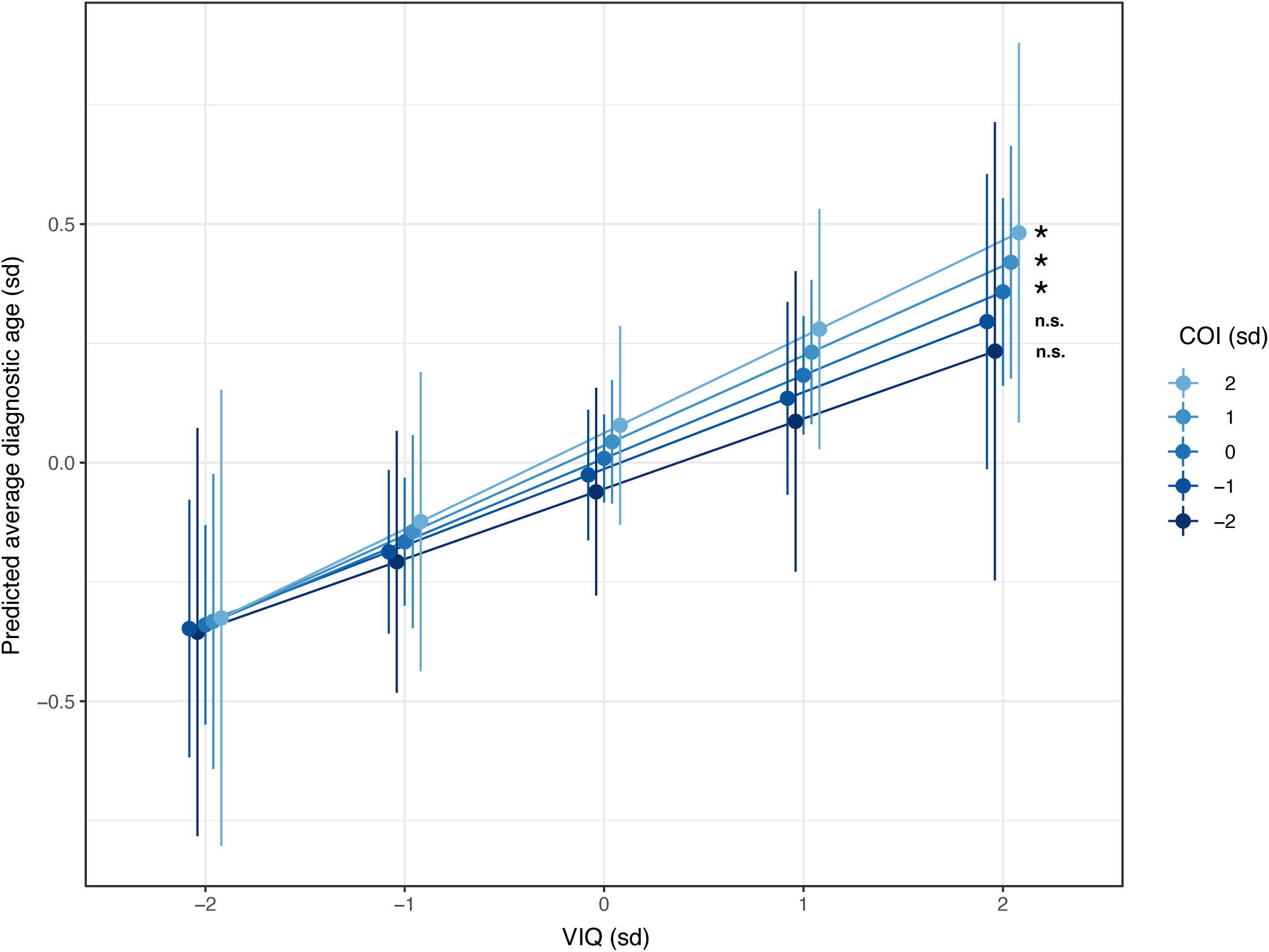
Predicted average diagnostic age in the full clinical sample at varying levels of VIQ and COI. Predicted average estimates of first age of autism diagnosis (y axis) within the full clinical sample were generated using the estimate_means tool from R’s modelbased package (Makowski et al., 2025). These estimates are plotted at representative values of verbal IQ (on the x axis) and COI (blue-shaded lines where darker=lower scores), with standard errors around the estimate indicated as vertical whiskers. All values depicted are normed and standardized such that zero indicates the sample mean and each one unit change represents a standard deviation (sd). Significance indicators (*: p > .05; n.s.: non-significant) on the slopes of the prediction lines were derived using the John-son-Neyman test.

**Table 5.** Model estimates in full clinical sample and clinical subsample.

| Coefficients | $\beta$ | B | SE | *t* | *p* |
| --- | --- | --- | --- | --- | --- |
| **Full Sample (N=641)** |  |  |  |  |  |
| *Reference sex: male* |  |  |  |  |  |
| Intercept | -- | -0.04 | 0.05 | -0.90 | .369 |
| VIQ | 0.14 | 0.14 | 0.05 | 2.99 | .003* |
| COI | 0.03 | 0.03 | 0.05 | 0.72 | .472 |
| Female | 0.04 | 0.10 | 0.09 | 1.07 | .284 |
| VIQ × COI | 0.10 | 0.10 | 0.04 | 2.16 | .031* |
| VIQ × Female | 0.04 | 0.07 | 0.09 | 0.74 | .458 |
| COI × Female | 0.00 | 0.00 | 0.10 | 0.04 | .972 |
| VIQ × COI × Female | -0.09 | -0.17 | 0.09 | -1.87 | .062† |
| RMSE: 0.98; Adj. $R^2$: 0.03; $F(7,633)=3.57$, $p < .001$ | | | | | |
| **Subsample (N=483)** |  |  |  |  |  |
| *Reference sex: male* |  |  |  |  |  |
| Intercept | -- | -0.06 | 0.05 | -1.14 | .254 |
| VIQ | 0.13 | 0.13 | 0.05 | 2.48 | .014* |
| COI | 0.01 | 0.01 | 0.05 | 0.20 | .844 |
| Female | 0.07 | 0.16 | 0.11 | 1.40 | .162 |
| VIQ × COI | 0.12 | 0.12 | 0.05 | 2.37 | .018* |
| VIQ × Female | 0.06 | 0.12 | 0.11 | 1.05 | .293 |
| COI × Female | 0.02 | 0.04 | 0.12 | 0.31 | .756 |
| VIQ × COI × Female | -0.13 | -0.26 | 0.10 | -2.47 | .014* |
| RMSE: 0.97; Adj. $R^2$: 0.04; $F(7,476)=3.53$, $p=.001$ | | | | | |

### Clinical Subsample

VIQ was also significantly correlated with COI in the clinical subsample (*r*(482)=0.22, *p* < .001); however, generalized variance inflation factors for the final model were all 1. The overall model was significant (*F*(7,476)=3.56, *p* < .001, Adj. *R*^2=^0.04; Table 5), as was the three-way interaction among assigned sex, COI, and VIQ (*B*=-0.26, *p*=.014). The VIQ (*B*=0.13, *p*=.014) and VIQ × COI (*B*=0.12, *p*=.018) terms were significant. These findings suggest that COI’s influence on the relationship between VIQ and diagnostic age functions differently for AFAB and AMAB individuals. Johnson-Neyman testing indicated that for AFAB individuals, COI significantly moderated the relationship between VIQ and diagnostic age only at lower values (COI < 0.28). By contrast, for AMAB, COI significantly moderated the relationship between VIQ and diagnostic age only at higher values (COI > -0.21; Figure 3).

**Figure 3.**
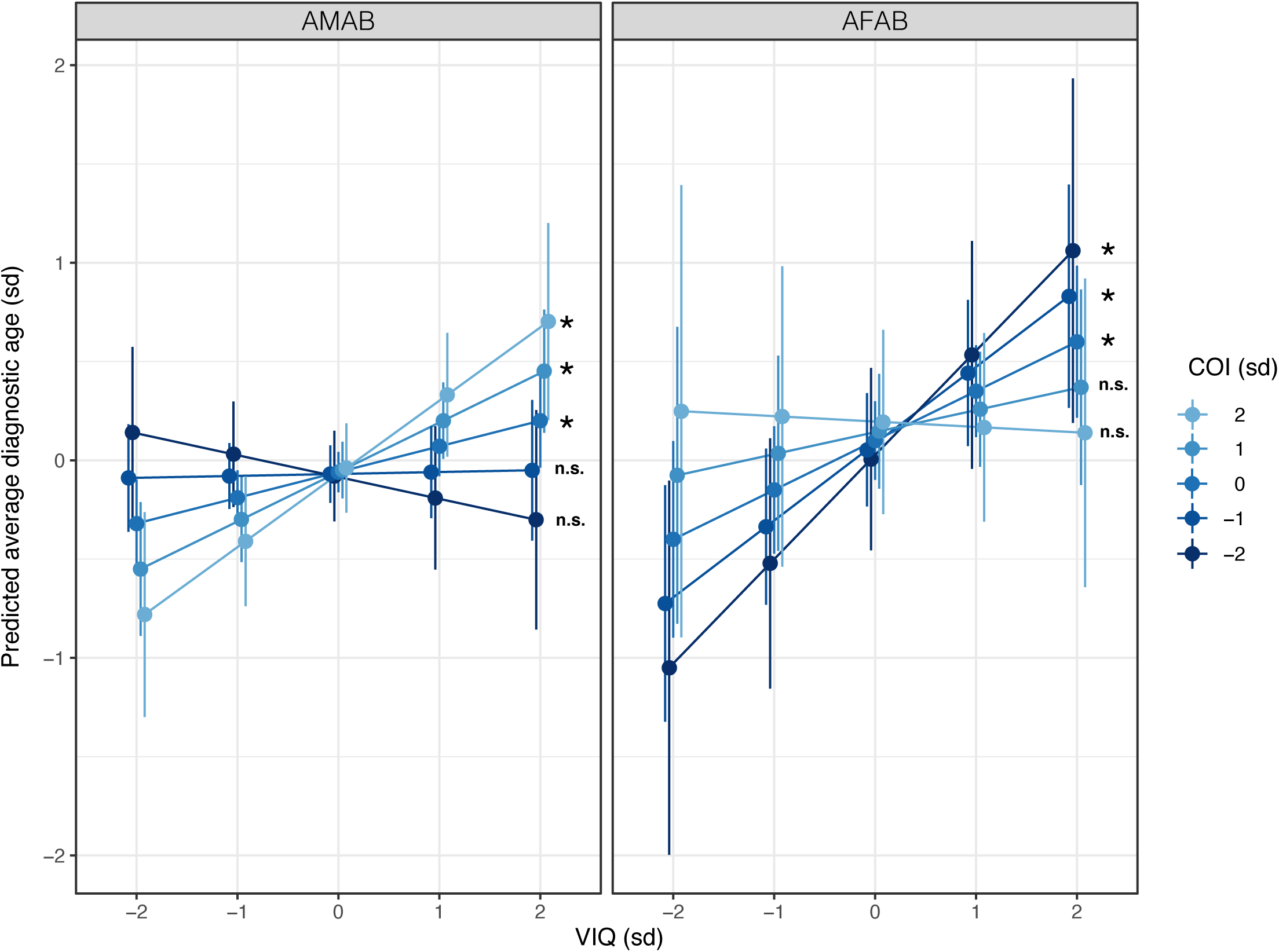
Predicted average diagnostic age in the clinical subsample by assigned sex at birth, at varying levels of VIQ and COI. Predicted average estimates of first age of autism diagnosis (y axis) within the clinical subsample were generated using the estimate_means tool from R’s modelbased package (Makowski et al., 2025). These estimates are paneled by assigned sex (AMAB: assigned male at birth; AFAB: assigned female at birth) and plotted at representative values of verbal IQ (on the x axis) and COI (blue-shaded lines where darker=lower scores), with standard errors around the estimate indicated as vertical whiskers. All values depicted are standardized such that zero indi-cates the sample mean and each one unit change represents a standard deviation (sd). Signifi-cance indicators (*: p > .05; n.s.: non-significant) on the slopes of the prediction lines were derived using the Johnson-Neyman test.

## Discussion

This study extends prior research investigating the intersection of multiple factors with sex assigned at birth in predicting diagnostic timing of autism. The present findings are novel in the use of the Childhood Opportunity Index as a comprehensive measure of neighborhood-level SDOH, encompassing multiple indicators of resources impacting childhood outcomes (i.e., education, health, and socioeconomic). An additional strength is the ability to compare two distinct samples: an assigned sex-balanced research cohort using stringent diagnostic confirmation and exclusion criteria, and a clinical sample in a demographically diverse metropolitan area using expert clinician judgment.

Overall, our results indicate that, consistent with prior literature, higher VIQ predicts later autism diagnosis. These results are observed both in a “clean” research sample with strict (but typical) inclusion/exclusion criteria, as well as a “real-world” clinical sample representing youth who are diverse in sociodemographic background and clinical presentation. The early identification literature indicates that caregiver worries related to language and communication are often the presenting concern for an autism diagnostic evaluation (e.g., Herlihy et al., 2015; Matheis et al., 2017). While lower verbal abilities tend to trigger evaluation, especially high verbal abilities may, on the other hand, tend to mask core autism features. This may be particularly true at younger ages where language abilities (expressive and receptive language; pre-literacy and literacy skills) dominate educational goals and social and adaptive expectations are still relatively simple.

Contrary to our hypothesis, assigned sex did not show evidence of a two-way interaction with VIQ in any of our samples; nor were there any main effects of sex. Rather, a more complex story emerged when we accounted for various SDOH via COI score. Within both the full clinical sample and the clinical subsample, there was a significant main effect of VIQ and VIQ × COI interaction. In the full sample, there was a trend toward a three-way interaction among sex, VIQ, and diagnostic age, and this became significant in the subsample. This emergence of a three-way interaction is likely attributable to the exclusion of participants with FSIQ<70 (as assigned sex differences in autism presentation are strongest in non-intellectually disabled samples), as well as to the removal of cases with co-occurring medical and genetic conditions (as these conditions may bring individuals to clinical attention earlier).

Among AFAB youth in this subsample, higher VIQ in the context of lower COI predicted a later age at diagnosis, while the opposite pattern held true for AMAB youth. Prior research has also found that girls/women with high VIQ are most likely to experience delays in autism diagnostic timing, which has been linked to a more subtle clinical presentation and higher rates of camouflaging (McQuaid et al., 2022). Our present findings suggest that AFAB youth who are living in low-resource areas and who have strong verbal abilities are the most vulnerable to delayed diagnosis of autism. In overburdened systems triaging access to limited resources, children with high VIQ, no other co-occurring medical conditions, and who exhibit a less “traditional” presentation of autism (as may be seen in many female cases) may be more likely to be overlooked.

Among AMAB youth in the subsample, higher VIQ predicted later age at diagnosis primarily for youth living in areas with average or higher resource levels, with the estimated association between VIQ and age at diagnosis relatively flat for those in under-resourced areas. This finding was unexpected, and appears to run counter to work indicating, for example, that children from families of higher socioeconomic status are more likely to receive developmental screening in a pediatric care setting than more disadvantaged peers (Hirai et al., 2018), and that boys are more likely to screen positive on the M-CHAT-R/F (an autism screening tool widely used in general pediatric care settings in the US) than girls (Eldeeb et al., 2023). As noted in the DSM-5 (APA, 2013), while autism’s neurodevelopmental characteristics will be present in the individual within the first two years of life, **observable** features sufficient to meet diagnostic criteria may not become evident until the demands of the context exceed the developing individual’s capacity to compensate or adapt. It may be that those demands differ dependent not only on resources but also assigned sex, leading to variability in when hallmark autistic traits “become visible” to clinical attention. Further research will be needed to understand these complex and sometimes counterintuitive patterns.

Overall, the results of this study indicate that both assigned sex at birth and SDOH, and the interaction of these factors with one another, are important in understanding diagnostic timing in autism. These factors likely also interact with individual, familial, and systemic factors more broadly to influence the autism diagnostic process.

These findings have important implications for understanding and designing future research investigating assigned sex differences in autism. As seen in the present study, research samples are likely not representative of the broad population of autistic people, due in part to potentially overly stringent case ascertainment approaches and restrictive exclusion criteria. While intended to ensure internal validity of findings through clear characterization, these restrictive criteria fail to capture true assigned sex differences, due to constraining variability of the sample (Mo et al., 2021). External validity is also threatened by samples that are not representative of the national population in terms of socioeconomic and ethnoracial diversity (Maye et al., 2022; Steinbrenner et al., 2022), as seen in the differences between our clinical and research samples. As has been previously found in both neurotypical (von Stumm & Plomin, 2015) and autistic (Ratto et al., 2021) populations, higher COI was associated with higher VIQ, indicating that even in the context of autism, SDOH has a powerful relationship with performance on VIQ measures. This underscores the importance of considering the role of environmental factors in clinical practice and research, and has important implications for policy to mitigate these risks.

Though not the focus on this study, it is notable that the average age at diagnosis in the clinical sample (∼9-10y) was substantially later than the age at diagnosis in the research sample (∼6 y), across both assigned sexes. Research studies intending to investigate sex and/or gender differences in autism, which seem to be more apparent later in development, may also need to consider diagnostic timing of their participants as a factor in recruitment. The clinical and research samples here likely represent distinct experiences of autism, due both to the differences in diagnostic timing and to factors that contributed to diagnostic delays in the clinical sample.

### Limitations

Each of our samples has unique strengths and limitations. By performing conceptually similar analyses across these samples, we sought to capitalize on the cohorts’ strengths while helping to control for their limitations. In the research cohort, strengths included an assigned sex-balanced sample and rigorous, consistent characterization across all participants; however, this sample was smaller, majority White, and lacked representation of individuals with intellectual disability, co-occurring medical conditions, and other complex presentations. Our inability to model SDOH factors in this sample was particularly notable. The limited available data on income and parental education from the research sample suggest that families with significant financial and educational resources were over-represented. This may have contributed to their ability to obtain relatively timely diagnoses. However, it is also possible, given the substantial missingness in the income data, that lower income families were more reluctant to share financial information with the researchers. Particularly given the known association between VIQ and socioeconomic status (Hackman & Farah, 2009; von Stumm & Plomin, 2015), it is possible that socioeconomic factors may have contributed to diagnostic age in this sample more than VIQ, or may have interacted with VIQ and sex assigned at birth as we observed in our clinical sample.

Strengths of the clinical cohort included its size and diversity in multiple dimensions (ethnoracial identity, socioeconomic status, medical status, IQ). However, given this cohort was characterized based on each individual’s clinical needs at the time of presentation, VIQ measures used varied; moreover, age at assessment and age at diagnosis were confounded.

The variability found in this cohort constitutes both a strength and a limitation. It is more representative of how cases typically present for diagnosis in the community than the “clean” research sample, with its many exclusion criteria; however, this variability, particularly in the measures used for phenotypic characterization, can undermine internal validity.

A major limitation of this study was its lack of characterization and modeling of gender in addition to assigned sex. Although participant gender was available in the clinical sample, it was not collected in the research sample. As gender and assigned sex at birth are distinct constructs, and gender diversity is elevated in autistic samples and associated with increased mental health risks, investigation of gender is an important priority for future research (Strang et al., 2023). Finally, although we investigated the interactive effects of VIQ and sex assigned at birth, investigation of the role of ethnoracial identity was beyond the scope of this paper. Given the known vulnerability to racial bias embedded in most IQ tests (Kaplan, 2015), and the conflation of ethnoracial identity and SDOH in the US, this bears further investigation, both independently and in the context of sex/gender differences. Though complex and not easily assessed, intersectionality is a critical factor at play in outcomes related to missed and delayed diagnosis. A full understanding of the impact assigned sex has on the experience of autism can only be attained when our samples are fully representative of the diversity of the autistic community (Diemer et al., 2022).

### Conclusions

Our results indicate that higher VIQ predicts later autism diagnosis across samples with varying autism ascertainment. Among those without co-occurring intellectual disability or significant known medical or genetic conditions, lower SDOH exacerbated the association between higher VIQ and later diagnosis for AFAB, while the opposite was true for AMAB. These findings suggest that careful assessment of SDOH and its interaction with assigned sex is necessary to fully understanding how assigned sex impacts age at autism diagnosis, and underscore the importance of comprehensive and nuanced autism diagnostic assessment.

## Data Availability

Raw phenotyping data from the research sample is available through the National Institutes of Health Data Archive (Collection #2021); a limited, de-identified version of the clinical dataset containing only information necessary to replicate analyses may be available upon request from the senior author.

## Acknowledgements

This work was funded by the National Institutes of Health (R01MH100028, MPIs: Pelphrey, Kenworthy, & Jack; U54HD090257, PI: Gallo; K01MH129622, PI: McQuaid) and the Clinical and Translational Science Institute at Children’s National (Award Grant No. UL1TR001876). The authors are grateful to the GENDAAR Consortium members for their contributions to data collection and management, which were utilized in this work. The opinions expressed in the manuscript are solely the authors’ own and do not necessarily represent those of the National Institutes of Health or US federal government.

## Ethical Considerations

### Research Sample

Procedures were approved and data collected under University of California Los Angeles Institutional Review Board (IRB) #10-000387, Boston Children’s Hospital IRB-P00004852, and Yale University Human Investigation Committee #1206010363. Parents provided informed written consent, and children provided written assent. *Clinical Sample.* The clinical sample was drawn from archival data collected through a continuing study approved and monitored by the Children’s National Institutional Review Board (IRB) #Pro00001320.

### Key points

- We used an assigned sex-balanced *research* sample and a diverse *clinical* sample to analyze the impacts of sex assigned at birth, verbal ability, and social drivers of health on diagnostic timing.
- Across samples with different inclusionary criteria and case ascertainment, stronger verbal abilities relate to later age at first autism diagnosis.
- In a diverse clinical sample, the relationship between stronger verbal abilities and later age at diagnosis was moderated by neighborhood social drivers of health, such that the relationship between verbal abilities and diagnostic timing did not depend on area resources for those in low-resourced areas.
- When cases of intellectual disability, syndromic autism, or significant medical complexity were removed from the clinical sample, sex emerged as a moderator of the interaction between neighborhood social drivers of health and verbal ability in predicting age of diagnosis, such that female youth from low-resourced areas received later diagnoses when they had higher verbal IQ, while male youth showed the opposite pattern.

## Supporting Information

### Data transformations

As described in the main text, R’s bestNormalize (Peterson, 2021) selected the best transformations for each continuous variable included in each model, using the Pearson chi-square test for the composite hypothesis of normality. Specifically, the transformation giving the lowest value for the Pearson P statistic divided by its degrees of freedom (estimated out-of-sample via cross-validation with 10 folds and 5 repeats) was used.

#### Research sample

For VIQ, the transformation selected and applied was the standardized square root (*M* before standardization [std.] = 10.07, *SD* before std. = 1.10; Pearson P/df = 1.13). For diagnostic age, the transformation selected and applied was the std. logarithm base 10 (*M* before std. = 0.69, *SD* before std. = 0.27; P/df = 1.15).

#### Full clinical sample

For VIQ, the transformation selected and applied was the std. Box-Cox (λ = 1.27; *M* before std. = 261.18, *SD* before std. = 69.98; Pearson P/df = 1.37); for COI, it was the std. Yeo-Johnson (λ = 1.00, *M* before std. = 59.68, *SD* before std. = 28.90; Pearson P/df = 2.56); for diagnostic age, it was the std. Yeo-Johnson (λ = 0.37, *M* before std. = 3.73, *SD* before std. = 0.89; Pearson P/df = 1.11).

#### Clinical subsample

For VIQ, the transformation selected and applied was the std. square root (*M* std. = 10.05, *SD* before std. = 0.91; Pearson P/df = 1.05); for COI, it was the std. Yeo-Johnson (λ = 1.09, *M* before std. = 83.68, *SD* before std. = 40.48; Pearson P/df = 2.10); for diagnostic age, it was the std. square root (*M* std. = 3.08, *SD* before std. = 0.63; Pearson P/df = 1.15).

**Table S1.** Available socioeconomic characteristics of the full GENDAAR sample, by assigned sex at birth.

| **Research Sample (N = 164)** |  |  |  |  |
| --- | --- | --- | --- | --- |
|  | Female<br>*n* = 71 | Male<br>*n* = 93 | Sex<br>Diff. |  |
| | *N* | *n* | $\chi^2$ | *p* |
| **Biological Mother Education** |  |  |  |  |
| Less than high school | 1 | 3 | -- | -- |
| High school degree | 6 | 6 | 0.00 | 1.00 |
| Some college | 12 | 20 | 2.00 | .212 |
| Associate's Degree | 2 | 6 | -- | -- |
| Bachelor's Degree | 20 | 24 | 0.36 | .651 |
| Some graduate work | 4 | 5 | -- | -- |
| Graduate Degree | 18 | 24 | 0.86 | .445 |
| Missing | 8 | 5 |  |  |
| **Household Income** |  |  |  |  |
| \$0-\$5,000 | 0 | 0 | -- | -- |
| \$5,001-\$10,000 | 0 | 1 | -- | -- |
| \$10,001-\$15,000 | 2 | 2 | -- | -- |
| \$15,001-\$25,000 | 1 | 2 | -- | -- |
| \$25,001-\$35,000 | 2 | 3 | | -- |
| \$35,001-\$50,000 | 2 | 8 | | |
| \$50,001-\$75,000 | 6 | 8 | | |
| \$75,001-\$100,000 | 6 | 10 | | |
| \$100,001-\$150,000 | 6 | 12 | 6.13 | .295 |
| > \$150,000 | 13 | 19 | | |
| Missing | 33 | 28 |  |  |

**Note:** Sex differences in observed counts are calculated using chi-squared tests with *p* value computed by Monte Carlo simulation using 10,000 replicates.

**Table S2.**
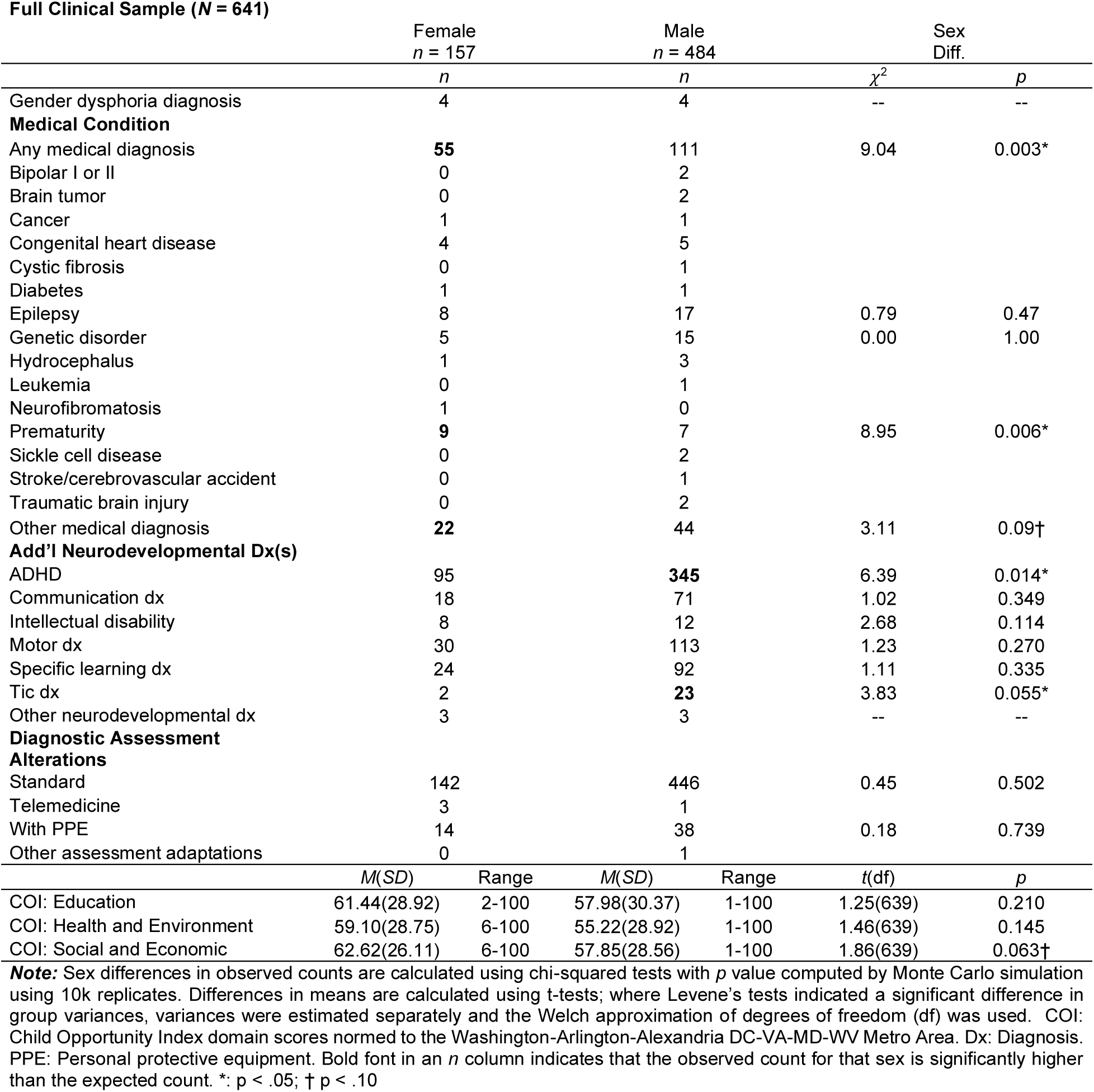
Additional descriptive characteristics of the full clinical sample, by assigned sex at birth.

**Table S3.**
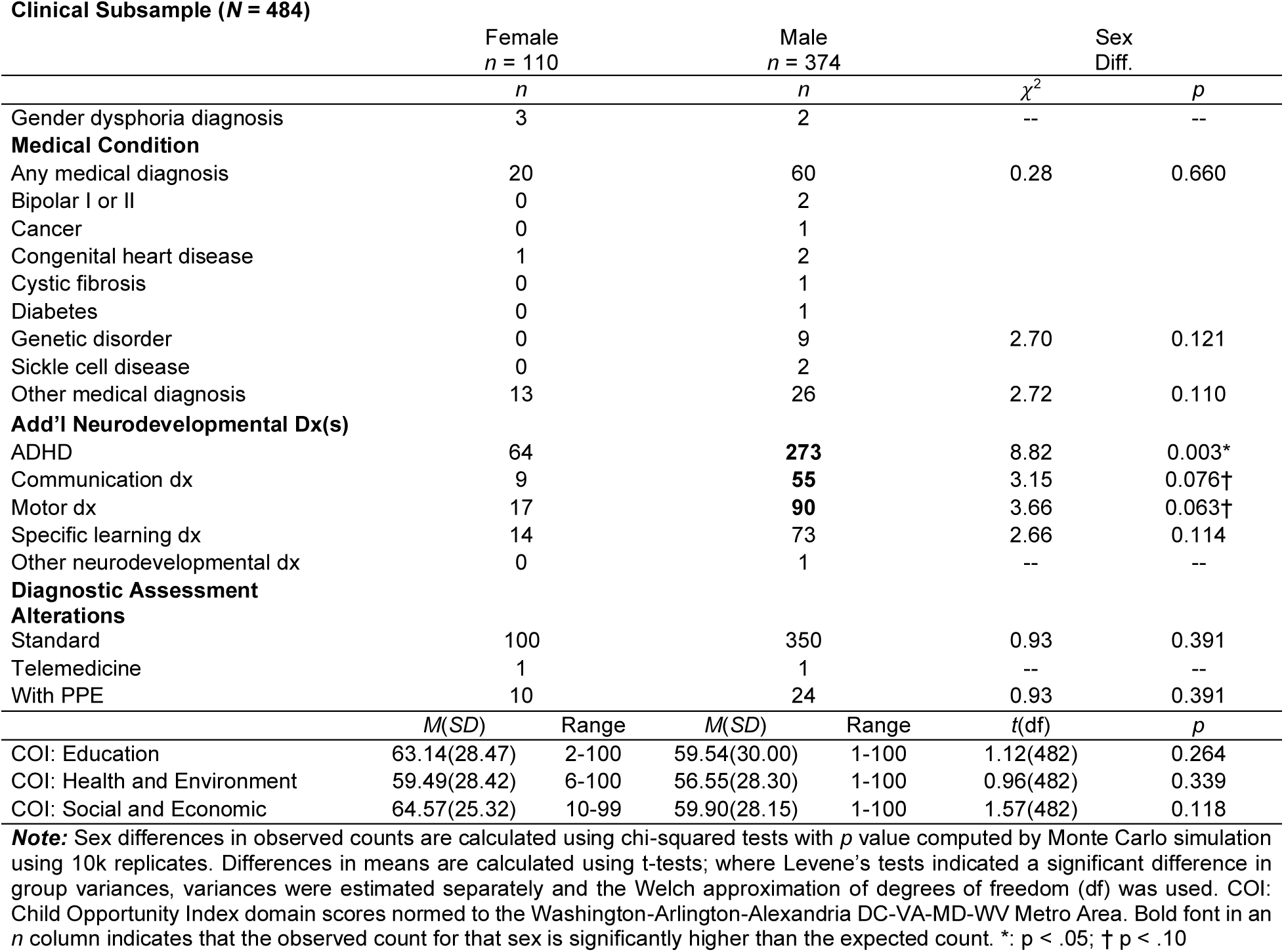
Additional descriptive characteristics of the clinical subsample, by assigned sex at birth.

| **Clinical Subsample (N = 484)** |  |  |  |  |  |  |
| --- | --- | --- | --- | --- | --- | --- |
|  | Female<br>*n* = 110 |  | Male<br>*n* = 374 |  | Sex<br>Diff. |  |
| | *n* | | *n* | | $\chi^2$ | *p* |
| Gender dysphoria diagnosis | 3 |  | 2 |  | -- | -- |
| **Medical Condition** |  |  |  |  |  |  |
| Any medical diagnosis | 20 |  | 60 |  | 0.28 | 0.660 |
| Bipolar I or II | 0 |  | 2 |  |  |  |
| Cancer | 0 |  | 1 |  |  |  |
| Congenital heart disease | 1 |  | 2 |  |  |  |
| Cystic fibrosis | 0 |  | 1 |  |  |  |
| Diabetes | 0 |  | 1 |  |  |  |
| Genetic disorder | 0 |  | 9 |  | 2.70 | 0.121 |
| Sickle cell disease | 0 |  | 2 |  |  |  |
| Other medical diagnosis | 13 |  | 26 |  | 2.72 | 0.110 |
| **Add'l Neurodevelopmental Dx(s)** |  |  |  |  |  |  |
| ADHD | 64 |  | **273** |  | 8.82 | 0.003* |
| Communication dx | 9 |  | **55** |  | 3.15 | 0.076† |
| Motor dx | 17 |  | **90** |  | 3.66 | 0.063† |
| Specific learning dx | 14 |  | 73 |  | 2.66 | 0.114 |
| Other neurodevelopmental dx | 0 |  | 1 |  | -- | -- |
| **Diagnostic Assessment** |  |  |  |  |  |  |
| **Alterations** |  |  |  |  |  |  |
| Standard | 100 |  | 350 |  | 0.93 | 0.391 |
| Telemedicine | 1 |  | 1 |  | -- | -- |
| With PPE | 10 |  | 24 |  | 0.93 | 0.391 |
|  | *M(SD)* | Range | *M(SD)* | Range | *t(df)* | *p* |
| COI: Education | 63.14(28.47) | 2-100 | 59.54(30.00) | 1-100 | 1.12(482) | 0.264 |
| COI: Health and Environment | 59.49(28.42) | 6-100 | 56.55(28.30) | 1-100 | 0.96(482) | 0.339 |
| COI: Social and Economic | 64.57(25.32) | 10-99 | 59.90(28.15) | 1-100 | 1.57(482) | 0.118 |

**Note:** Sex differences in observed counts are calculated using chi-squared tests with *p* value computed by Monte Carlo simulation using 10k replicates. Differences in means are calculated using t-tests; where Levene's tests indicated a significant difference in group variances, variances were estimated separately and the Welch approximation of degrees of freedom (df) was used. COI: Child Opportunity Index domain scores normed to the Washington-Arlington-Alexandria DC-VA-MD-WV Metro Area. Bold font in an *n* column indicates that the observed count for that sex is significantly higher than the expected count. \*: $p < .05$ ; † $p < .10$

## Abbreviations

AFAB: Assigned female at birth
AMAB: Assigned male at birth
COI: Childhood Opportunity Index
SDOH: Social drivers of health.

## Footnotes

1 This manuscript uses both identity-first and person-first language, in recognition of the multiple perspectives within autistic and autism communities on this issue.

